# Protocol for the AutoRayValid-RBknee Study: a Retrospective, Multicenter, Fully-crossed, Multi-reader, Multi-case Study Investigating the Effect of a Knee Osteoarthritis Severity Classification Model on Reader Diagnostic Accuracy

**DOI:** 10.1101/2022.08.29.22279328

**Authors:** Mathias Willadsen Brejnebøl, Liv Egnell, Michael Lundemann, Anders Lenskjold, Janus Uhd Nybing, Huib Ruitenbeek, Katharina Ziegeler, Jacob Visser, Kay Geert A. Hermann, Edwin H.G. Oei, Mikael Boesen

**Affiliations:** Department of Radiology, Frederiksberg and Bispebjerg Hospital, Copenhagen, Denmark; Radiobotics ApS, Copenhagen, Denmark; Department of Radiology, Charité - Universitätsmedizin Berlin, Berlin, Germany; Department of Radiology & Nuclear Medicine, Erasmus Medical Center, Rotterdam, The Netherlands

## Abstract

**Background:** Radiographic evaluation of knee osteoarthritis (KOA) commonly supports clinical findings. Ground truth is difficult to establish and concerns exist on the inter-and intrarater agreement of the findings. RBknee™ is a CE-marked and FDA-cleared AI tool for automatic assessment and reporting of radiographic KOA on standard projection radiographs.

**Objectives:** To investigate how the use of an AI tool affects the accuracy among human readers across three European hospitals in grading the severity of osteoarthritis and associated individual radiographic features. In addition, the performance of the AI tool will also be compared to reference standards established by experts in a stand-alone validation.

**Methods:** In this retrospective multicenter, fully-crossed, multi-reader, multi-case (MRMC) study, the AI support tool RBknee is introduced as a diagnostic intervention. Four Index Readers from each site (two orthopaedic surgeons and two radiologists) will read all studies twice in two runs separated by a washout period of at least four weeks. In both runs, the experiment will be arranged so that the AI-aid will be available for half of the images in the first session and for the second half of the images in the second session. The order of the images will be randomised in order to minimise temporal effects and biases. The primary endpoint is the difference in diagnostic test accuracy for radiographic KOA grading without and with the aid of the AI tool and will be measured as the ordinal weighted accuracy.

**Data:** The data includes radiographic images from 225 studies (unique patients, retrospective data) with weight-bearing bilateral PA/AP and LAT projections of the symptomatic knee(s). Each site contributes to the cohort with 75 studies of which 70 will be consecutive and 5 will be selected to balance the prevalence of radiographic KOA severity.

**Reference standard:** The reference standard will be established based on independent grading by three KOA Reference Experts and adjudicated by majority vote. Where impossible to resolve by majority voting, adjudication will be established by consensus.

**Index test, AI tool (stand-alone validation):** The diagnostic accuracy of RBknee will be tested against the reference standard.

**Index test, Index Readers:** The 12 readers will grade KL on the PA/AP projection and patellar osteophytes on the lateral projection.

**Administrative information:** *Title:* The trial is titled “AutoRayValid-RBknee”.

*Protocol version:* **Revision History** 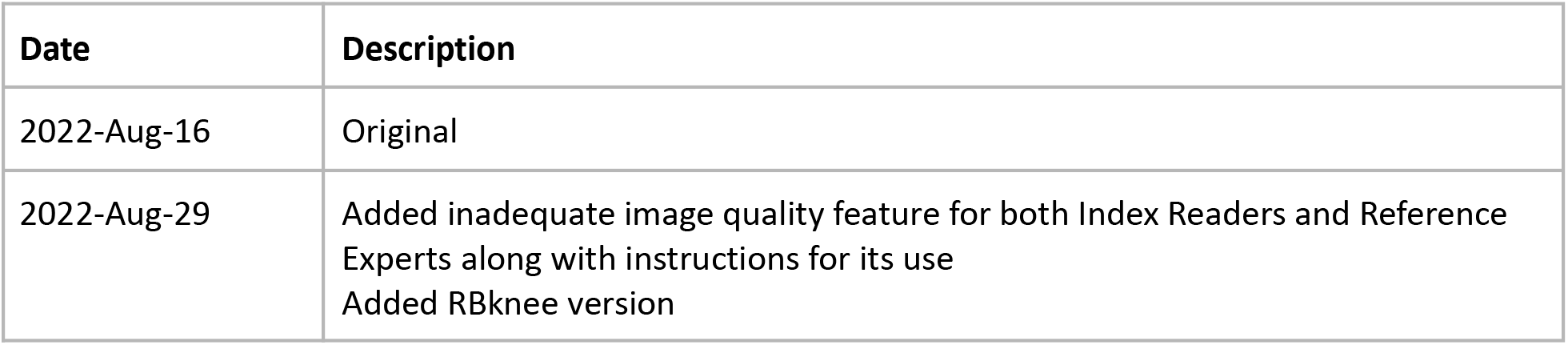

*Funding:* This project has received funding from the European Union’s Horizon 2020 research and innovation programme under grant agreement No 954221 for the EIC SME Instrument project AutoRay. The work only reflects the authors’ view and the European Commission is not responsible for any use that may be made from the information it contains.

**Roles and responsibilities:** *Authors’ contributions:* MWB, MB, EHGO, JV, and KGH initiated the study design and AL, JUN, KZ and HR helped with implementation. MWB, LE and MJL provided statistical expertise in clinical study design and MWB is conducting the primary statistical analysis. MWB, MJL and LE prepared the manuscript and all authors contributed to its refinement and approved the final manuscript.

*Sponsor Contact information:* **Trial Sponsor:** Radiobotics ApS **Contact name:** Liv Egnell **Address:** Esplanaden 8C, 1263 Copenhagen K, Denmark **Email:**

## Introduction

### Background and rationale

Knee Osteoarthritis (KOA) is the most common joint disease and is a major cause of disability in men and women worldwide. Twenty-three percent of the world’s population above 40 years as estimated by Cui et al., suffer from joint related problems attributable to KOA ^1^. Risk factors for KOA include older age, sex (female), overweight and positive family history. Osteoarthritis typically presents with a slow progression and can often be effectively managed conservatively with education, information access, exercise, weight loss and pharmacologically with analgesics, such as paracetamol and NSAID. However, patients are affected by varying degrees of disability and, as the disease Osteoarthritis of the knee is diagnosed clinically if the patient meets the following three criteria developed by the European League Against Rheumatism (EULAR) ^2^: (1) age ≥ 45 years, (2) has activity-related joint pain, and (3) does not suffer from morning-related stiffness or has morning stiffness that lasts no longer than 30 minutes. Radiography is recommended as a supplement in patients with clinical symptoms that are either younger than 45 years or if their pain is resistant to conservative therapy. The main radiographic findings associated with KOA are narrowing of joint space due to cartilage loss, osteophytes and several changes in the subchondral bone, such as sclerosis, cysts, shape changes and loss of bone volume ^2^.

The most commonly used grade for radiographic KOA classification is the Kellgren-Lawrence (KL) grading system, which was developed by Kellgren and Lawrence in 1957 ^3^. It is separated into five non-equidistant grades of increasing severity (0-4), where KL grade 2 or above is usually defined as radiographic KOA. The system has limitations and is associated with only-moderate reliability both in terms of intra-and interrater agreement ^4–6^. The Osteoarthritis Research Society International (OARSI) developed an atlas to classify the severity of each of the radiographic findings on ordinal scales ranging from 0 to 3 with increasing severity ^7^.

As life expectancy continues to increase the prevalence of KOA is expected to increase even further, resulting in a heavier workload on general practitioners and radiology departments. In addition, the radiological interpretation and scoring is subject to a great deal of unintended variation due to subjective interpretation of images often exacerbated by overly busy working environments among the reporting personnel such as radiologists and diagnostic radiographers. Reduced variation in image interpretation and more robust quantification of KOA will allow a more consistent diagnosis and open possibilities for earlier identification of radiographic markers of the disease as well as more exact tracking of KOA progression.

Computer assisted diagnosis systems have shown promising results in KL-grading of KOA, with performance similar to within-domain experts ^8–10^ and improving the reliability of human readers, when used as decision support ^11^. However, the aforementioned studies all rely on the same open source datasets or small clinical samples, limiting the generalizability of these results.

RBknee™is a software program with algorithms for automatic assessment and reporting of radiographic KOA on standard projection radiographs. Its standalone performance has been assessed using two large, publicly available research datasets ^12^ and on clinical data with radiologic readers of various experience levels ^13^. However, the impact on human reader accuracy and reliability when using RBknee as decision support on clinically acquired imaging data remains unknown.

### Trial aim

This trial aims to investigate how the use of RBknee affects the diagnostic accuracy among human readers across three different European hospitals and to estimate the diagnostic accuracy of RBknee for characterising the degree of knee osteoarthritis on plain X-ray.

## Objectives

The specific objectives are:

1. to assess how decision support from RBknee affects the diagnostic test accuracy and inter-and intrarater agreement of radiographic KOA classification using the KL-grade and a clinical relevance allocation on posterior-anterior/anterior-posterior knee radiographs among human readers of varying experience and specialties
2. to assess how decision support from RBknee affects the diagnostic test accuracy and inter-and intrarater agreement of binary classification of the presence of osteophytes proximally and distally on the patella on lateral knee radiographs among human readers of varying experience and specialties
3. to assess the diagnostic test accuracy of the RBknee tool for radiographic KOA classification on both the KL-grade and a subset of the OARSI-grades

### Hypotheses

The **primary null hypothesis** is that decision support from RBknee does not improve the diagnostic test accuracy of readers when grading radiographic KOA. The **primary alternative hypothesis** is that the diagnostic test accuracy of readers when grading radiographic KOA is improved when they receive decision support from RBknee.

The **secondary null hypothesis** is that KOA grading agreement among readers is not improved by decision support from RBknee. The corresponding **alternative secondary hypothesis** is that the agreement for KOA grading increases among readers when receiving decision support from RBknee.

### Trial design

The trial design is a multicenter, fully-crossed multi-reader, multi-case (MRMC) trial, with RBknee as a diagnostic intervention. The trial design is in accordance with the *Guidelines for clinical trial protocols for interventions involving artificial intelligence: the SPIRIT-AI extension* (SPIRIT-AI) ^14^, with reference to the *STARD 2015 guidelines for reporting diagnostic accuracy studies: explanation and elaboration* ^15^ and *Guidelines for Reporting Reliability and Agreement Studies* ^16^. It was further inspired by *Multireader Diagnostic Accuracy Imaging Studies: Fundamentals of Design and Analysis*^*17*^

## Methods: Participants, Intervention and Outcomes

### Trial setting

The trial is a collaboration between Radiobotics ApS and three European clinical Sites;

- Department of Radiology, Bispebjerg and Frederiksberg Hospital, Copenhagen, Denmark (BFH),
- Department of Radiology, Charité - Universitätsmedizin Berlin, Berlin, Germany (CUB) and
- Department of Radiology & Nuclear Medicine, and Erasmus Medical Center, Rotterdam, The Netherlands (EMC).

### Trial Sample

The Trial Sample will consist of patients referred for radiography of the knee on suspicion of KOA without acute-onset of current symptoms. The trial participants will be collated consecutively from the Picture and Archiving System (PACS) at the three hospitals in a retrospective manner.

### Eligibility criteria

The inclusion criteria at the patient level

- adults aged 20 years or older
- clinical suspicion of KOA without acute-onset of current symptoms

The exclusion criteria at the patient level

- knee arthroplasty or other foreign objects in close proximity to the knee joint
- study is from a patient for whom a study has already been included in his trial

The inclusion criteria at the data level

- radiographs obtained according to local protocols, with a **minimum of** one weight-bearing image of each knee in a ‘frontal’ view (PA or AP) and one lateral view of the symptomatic knee

The exclusion criteria at the data level

- images not of PA, AP or lateral projections
- signs of previous surgery

### Data extraction / sampling

At each site, studies of eligible patients will be consecutively extracted from the PACS until 150 studies have been collected as a Site Batch. A suitably trained person will perform the data extraction and initial screening according to eligibility criteria. This person cannot be a participant in the subsequent reading phase.

The following demographic and clinical variables are extracted and each site will note the study date where consecutive inclusion began and ended:

#### Study information

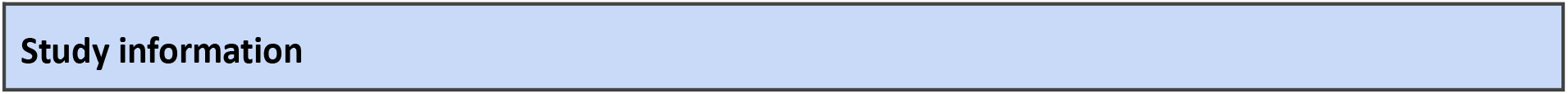
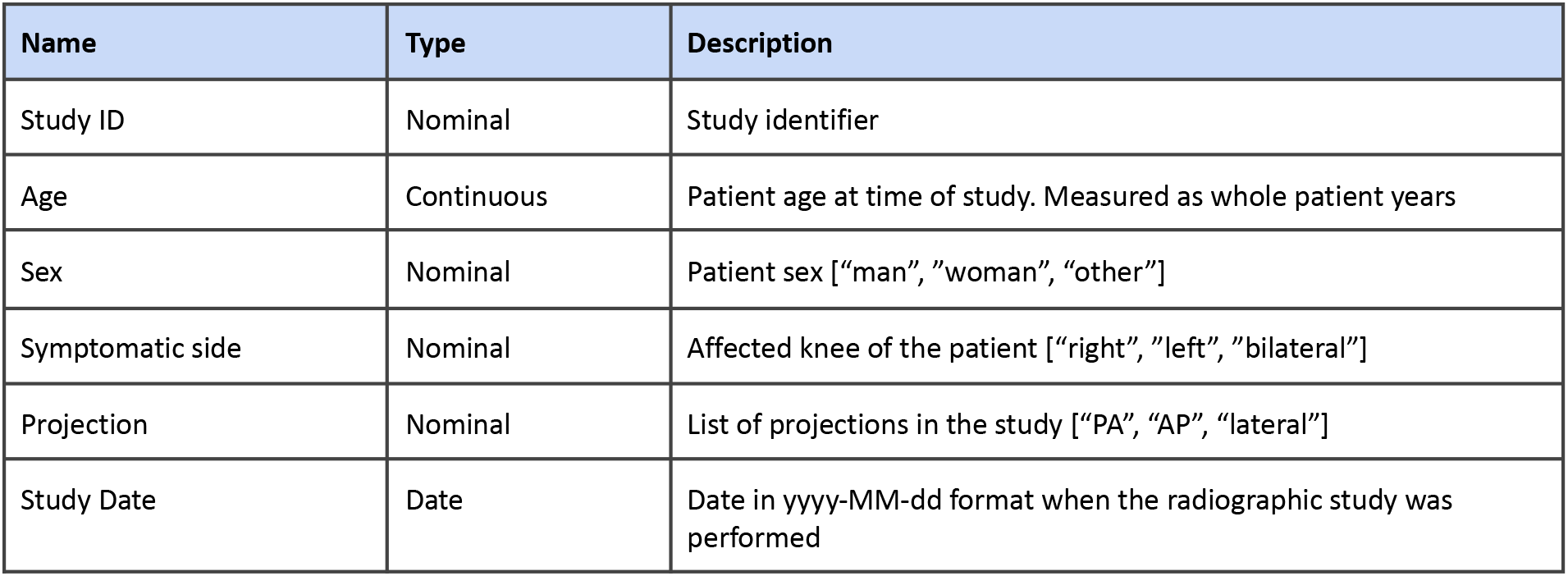

| Name | Type | Description |
| --- | --- | --- |
| Study ID | Nominal | Study identifier |
| Age | Continuous | Patient age at time of study. Measured as whole patient years |
| Sex | Nominal | Patient sex ["man", "woman", "other"] |
| Symptomatic side | Nominal | Affected knee of the patient ["right", "left", "bilateral"] |
| Projection | Nominal | List of projections in the study ["PA", "AP", "lateral"] |
| Study Date | Date | Date in yyyy-MM-dd format when the radiographic study was performed |

The studies are anonymized at each site and then encrypted and transported digitally to BFH. At BFH, a suitably trained person will include studies from each Site Batch into the final Trial Sample. From each Site Batch 70 studies will be included consecutively, again ensuring that eligibility criteria are met. The person will then include five additional studies from each Site Batch, ensuring that these studies are without signs of radiographic KOA. This will result in a Trial Sample of 70 + 5 + 70 + 5 + 70 + 5 = 225 radiographic studies of the knee.

### Endpoints

#### Primary

The primary endpoint is the difference in diagnostic test accuracy for radiographic KOA grading without and with the aid of RBknee. The diagnostic test accuracy will be measured as the ordinal weighted accuracy described by Obuchowski^18^ for assessing outcomes on an ordinal scale.

#### Secondary

The secondary endpoints are:

- the difference in accuracy between RBknee and Index Readers without decision support.
  - Metrics for clinical relevance and KL grading: ordinal weighted accuracy
  - Metrics for patellar osteophytes on the lateral images: binary balanced accuracy.

- the difference in agreement among Index Readers without and with decision support from RBknee
  - Metrics for clinical relevance and KL-grading: Quadratic weighted Light’s Kappa
  - Metrics for patellar osteophytes on the lateral images: Unweighted Light’s Kappa

#### Exploratory

The exploratory endpoints are:

- the diagnostic test accuracy of RBknee across features compared to the Reference test
  - Metrics for KL-grading: OARSI-JSN, OARSI-osteophytes on PA/AP: Ordinal weighted accuracy
  - Metrics for OARSI-subchondral sclerosis and patellar osteophytes on the lateral images: Binary balanced accuracy

- sensitivity, specificity, and F1-score will be reported for the items in the primary, secondary, and exploratory endpoints in addition to
  - For the dichotomized groups “No radiographic knee osteoarthritis” (KL 0-1) vs “Radiographic knee osteoarthritis present” (KL 2-3) and “Not end-stage radiographic osteoarthritis present” (KL 0-3) vs “End-stage radiographic osteoarthritis present” (KL 4)
- subgroup analysis among Index Reader specialty and experience
- description of the diagnostic accuracy of RBknee among relevant patient subgroups (sex and age)

### Sample size

The effect size for improving clinical relevance allocation (defined more thoroughly in the section Allocation to Clinical Relevance) “No radiographic knee osteoarthritis” (KL-grades 0-1), “Radiographic knee osteoarthritis present” (KL-grades 2-3), “End-stage radiographic osteoarthritis present” (KL-grade 4) is set at 5% in ordinal weighted accuracy. Assumptions for the power calculation were based on a preliminary dataset from BFH, and a biostatistician was consulted to arrive at the chosen methodology.

In this dataset, 50 posterior-anterior images of weight-bearing bilateral knees were KL-graded by six readers: two consultant radiologists, two reporting technologists, and two resident radiologists. All readers graded the images twice with a washout of four weeks between reading sessions. In addition, the two consultants had a consensus session where they arrived at consensus on all studies they had previously disagreed on, resulting in a consensus grade. RBknee also analysed all studies. Thus, for each study there is a baseline and a retest KL-grade for all six readers as well as a single consensus grade and a single RBknee grade. These KL-grades were allocated into the three categories mentioned above.

Synthetic data were used to determine the required sample size. A synthetic dataset with a sample size *n* was created by resampling the data in the original dataset and introducing noise (simulating intra-and interrater variance) based on the seniority of readers. This variance was added to the baseline and the retest grades in the dataset: The probabilities of KL-grade shifting one grade up or down was set at 0.05, 0.10 and 0.15 for consultants, reporting technologists and residents, respectively.

To simulate RBknee decision support, the probability for readers aided by the RBknee support to change the KL-grade toward the grade outputted by RBknee was assumed to be 0.05, regardless of seniority.

Permutation tests for ordinal weighted accuracy both without and with RBknee support (see dataset creation above) were performed. The total number of studies, *n*, was varied until a statistical difference between the two distributions could be consistently identified (p<0.05). A total number of 225 studies from 225 unique patients, 75 from each clinical site, will be included in this trial.

### Recruitment

Not applicable. The trial uses retrospective data and will be conducted in accordance with the Declaration of Helsinki ^19^ and the General Data Protection Regulation (GDPR) ^20^. Prior to the start of any study activities at each of the sites, local ethics committee opinion and approval from national competent authorities will be collected and if deemed required informed consent will be collected prior to inclusion of patients.

## Methods: Assignment of Interventions, Features and Readers

### The Kellgren-Lawrence grade

On the PA/AP radiograph, readers are requested to grade each knee according to the Kellgren-Lawrence grade^3^, with

1. no OA: no OA features
2. doubtful OA: possible osteophytic lipping, doubtful joint space narrowing
3. mild OA: definite osteophytes, possible joint space narrowing
4. moderate OA: moderate multiple osteophytes, definite joint space narrowing, some sclerosis, possible bone contour deformity
5. severe OA: large osteophytes, marked joint space narrowing, severe sclerosis, definite bony contour deformity

### Allocation to Clinical Relevance

Three clinically relevant groups are identified based on the KL scale:

“No radiographic knee osteoarthritis”: represents KL-grades 0-1 and a clinical heuristic that there is no certain evidence of radiographic osteoarthritis. In the clinic, this may warrant further workup.

“Radiographic knee osteoarthritis present”: represents KL-grades 2-3, and a clinical heuristic that there is radiographic evidence of osteoarthritis. According to many clinical guidelines, no further imaging workup is required at this point. Usually, there is no radiographic evidence for referral for further evaluation by an orthopedic surgeon unless other clinical information strongly suggests this cause of action.

“End-stage radiographic osteoarthritis present”: represents KL-grade 4, and clinical heuristic that there is complete or almost complete destruction of the joint. There is radiographic evidence that supports the patient being referred for evaluation by an orthopedic surgeon if other clinical information also suggests this cause of action.

Furthermore, the KL-grade may also be dichotomized into absence/presence of radiographic tibiofemoral OA and will be defined as “KOA not present” (KL < 2) and “KOA present” (KL >= 2).

### Choice of Knee Laterality

Posterior-anterior, weight-bearing radiographs of the knee are usually performed on both knees simultaneously. To avoid intra-patient correlation, only one of the knees will be used in the trial. If information on the symptomatic side is present and it is unilateral, that side is chosen for this trial. Otherwise, the chosen laterality will be determined randomly using a 0 to 1 random number generator with values less than 0.5 being the left knee and otherwise the right knee (using the random core module in Python 3.9.5).

### Training of readers

Prior to the first reading session, each reader will receive training in the use of the annotation platform ANOVI and on how to interpret the output provided by RBknee. This will allow the readers to get acquainted with the annotation platform ANOVI workflow and ensure the correct use of the RBknee output. An example of the ANOVI DICOM-viewer is demonstrated in Figure 1 and a filled scoring panel is demonstrated in Figure 2.

**Figure 1.**
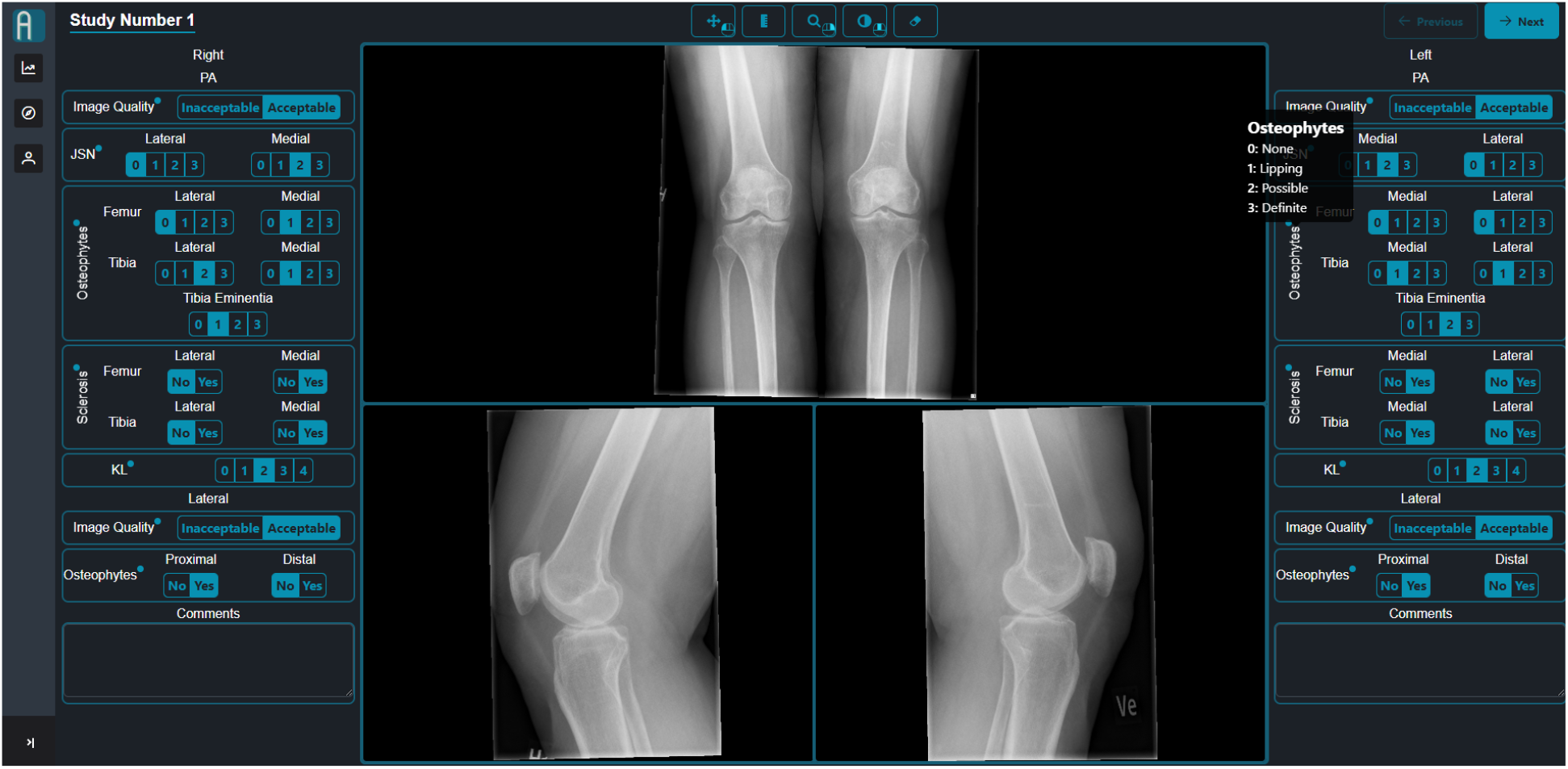
User interface and scoring panel in the annotation platform ANOVI (Reference Expert’s view).

**Figure 2.**
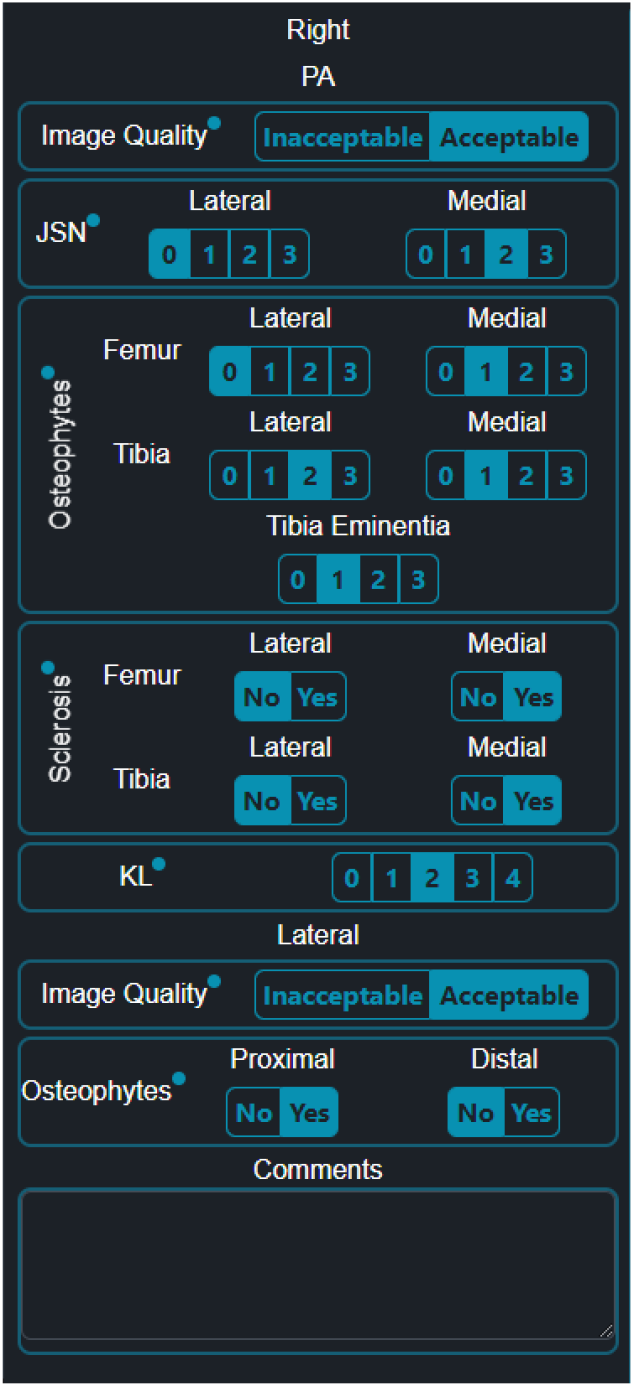
Dedicated scoring panel for AutoRayValid-RBknee in ANOVI (Reference Expert’s view).

**Figure 3.**
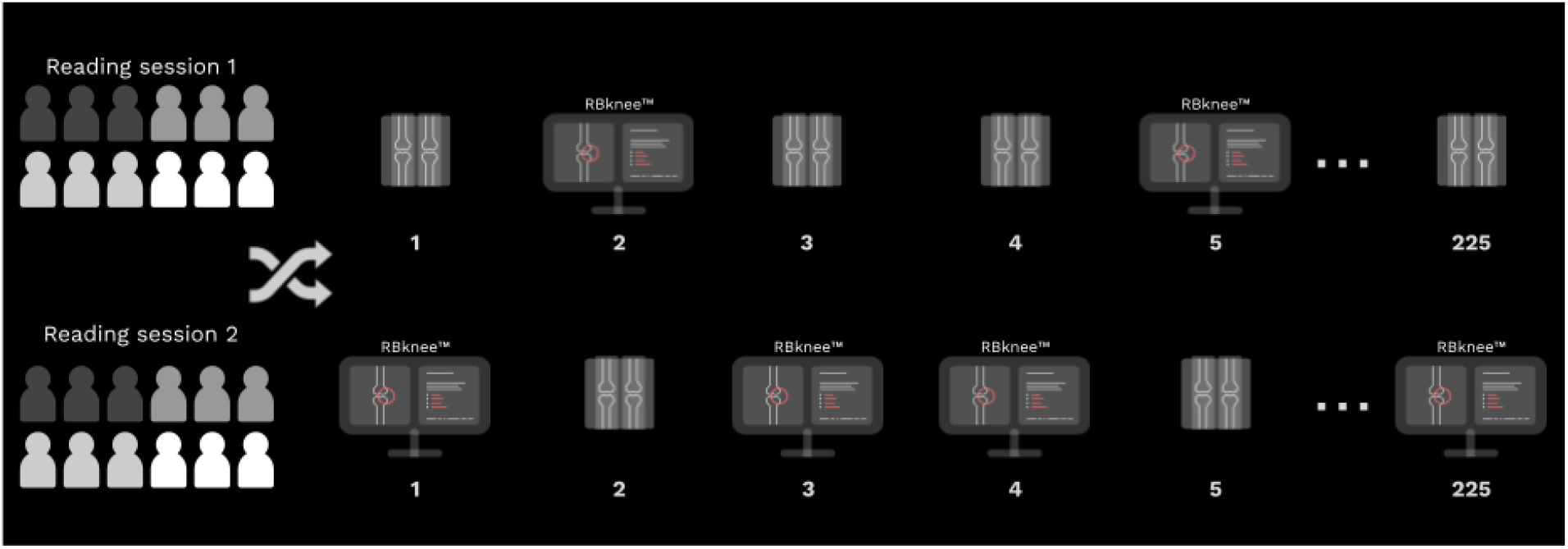
Paired reader trial, without and with decision support. Readings of the studies are done twice, in two separate sessions. For each study, RBknee will be available in one of the sessions and the order of which RBknee will be available is randomised between the sessions.

**Figure 4.**
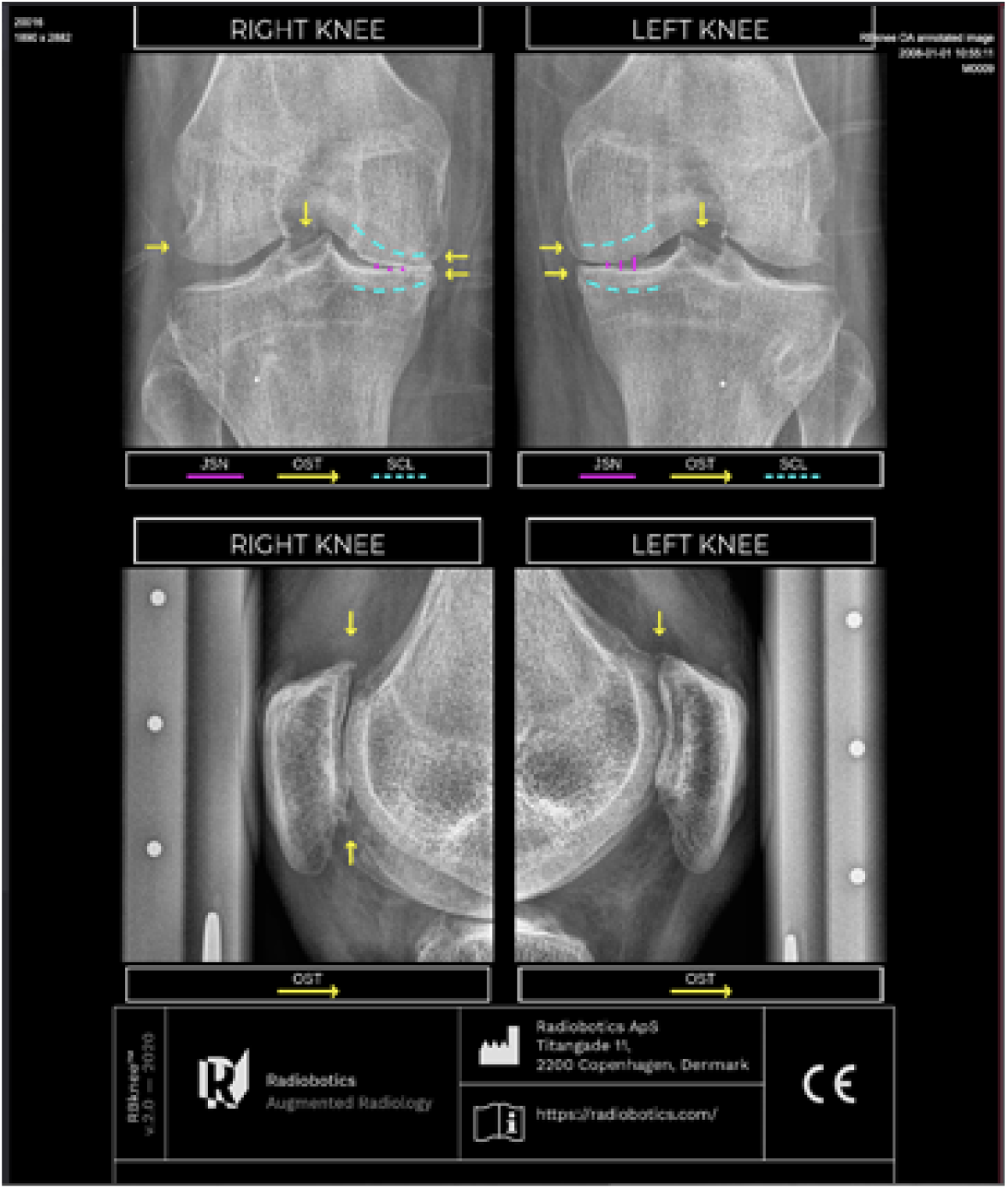
Illustration of output from RBknee.

**Figure 5.**
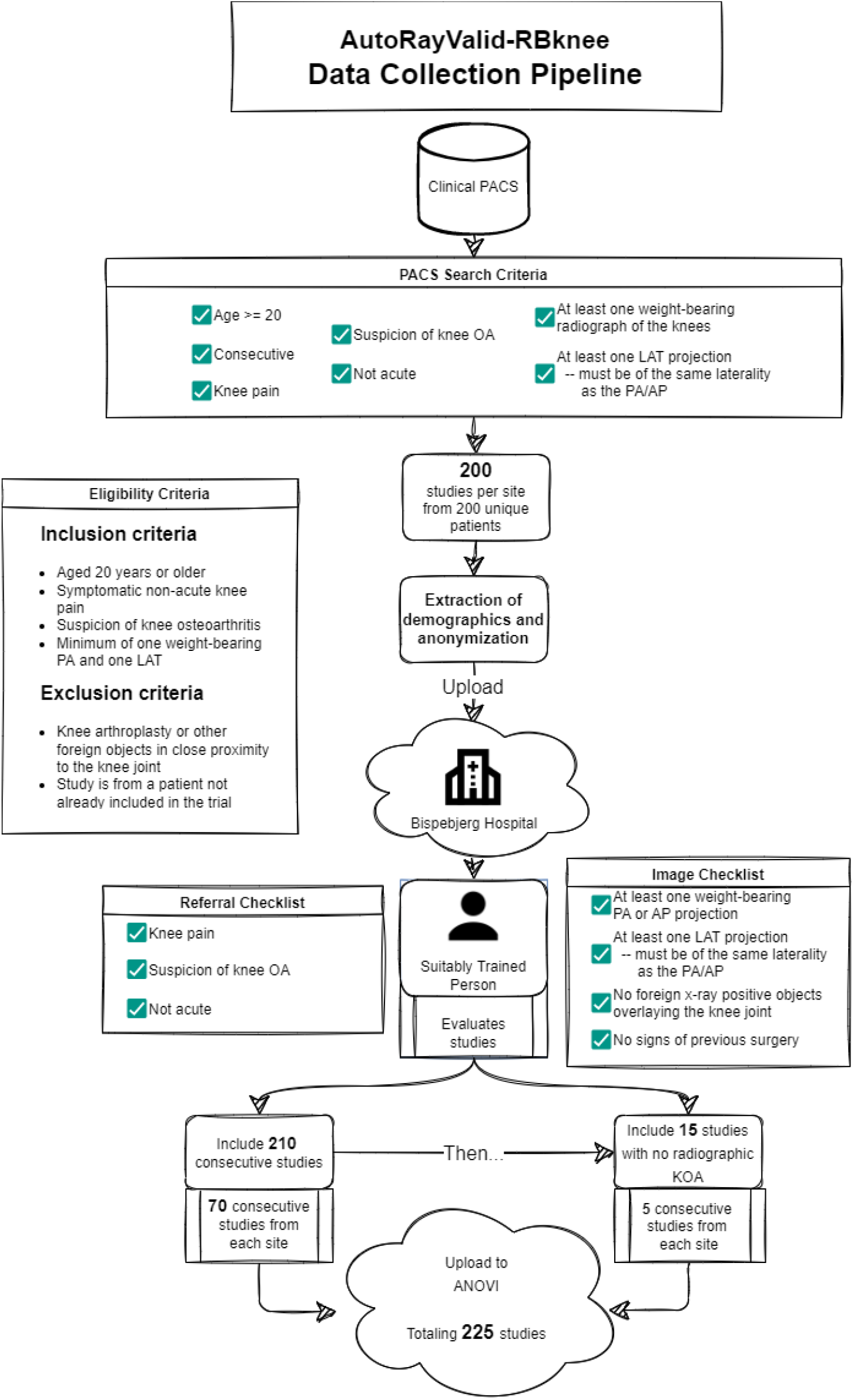
Data Collection Pipeline

### Reference test: Reference Experts

Each of the three clinical Sites will recruit one KOA Expert that will be used to construct a Reference Atlas (see later) and establish the Reference Standard. The KOA Experts are defined as individuals with extensive (>10 years) experience with clinical KOA reporting and research experience with KL and OARSI grading. Before the grading of trial studies begin the Reference Experts will convene in three separate Alignment Sessions. During these Sessions the Reference Experts will thoroughly discuss how they evaluate each separate feature mentioned below. Furthermore, the Reference Experts will create a Reference Atlas during these sessions (using a separate dataset from BFH) which will be made available to the Index Readers during trial readings.

The three Reference Experts will grade all of the 225 studies **once** without any decision aid.

#### Features

For each knee on the PA/AP projection, the Reference Experts will assign one KL-grade (0-4) as well as OARSI grades for medial and lateral joint space narrowing (0-3), osteophytes (0-3) and rate subchondral sclerosis (no/yes) on the medial and lateral distal femur and proximal tibia. In addition, the Reference Experts will rate osteophytes on the tibial eminence (no/yes). They will comment on any additional findings using a free-text field.

For the lateral projection, the Reference Experts will rate osteophytes (no/yes) on the proximal and distal patella. They will comment on any additional findings using a free-text field.

#### Assignment of final reference grade

The reference standard will be based on majority voting. For cases where majority voting would leave an invalid result, the three Reference Experts will convene and, based on discussion, will adjudicate a final grade. The Reference Experts are blinded to the output of RBknee and the other Index Readers.

### Index test: Index Readers

It is expected that readers are accustomed to reading and grading radiographic KOA in clinical practice. However, prior experience with KL-grading or the OARSI atlas is not a prerequisite. Prior to the Index Reader Sessions, an atlas of radiographic KOA will be developed by the three Reference Experts which will be available to all Index Readers during all their readings. No images from this atlas are present in the Trial Sample. Index Readers will be instructed that their assigned KL-grades should correspond to the Allocation to Clinical Relevance section as described above.

#### Interventions

The trial uses retrospective data; hence no patient will undergo a supplementary examination or radiation exposure, nor will it influence their treatment.

RBknee (v2.1) will be the diagnostic intervention.

Randomisation to the intervention will be applied using a random number generator (via the random core package in Python v. 3.9.5) so that for half of the patient studies, the aid will be present at the first read and vice versa. Randomisation will be performed by a suitably trained person, performing the data management.

#### First reading session

Readings will be done in the imaging trial platform; *ANOVI* (**DICOM-viewer with dedicated scoring panel**). For half of the patient studies, only the original radiographs will be available to the reader. For the other half of the patients, the original radiographs will be available together with the output from RBknee. Readers are blinded to the study indication, original study report, results of the reference test, and the results of all other index tests.

#### Second reading session

The second reading session will be similar to the first reading session, except that the intervention is reversed, i.e. patient studies that were read without aid in the first session will be read with aid in the second session and vice versa.

The second session will be separated in time by a wash-out period of at least four weeks. The readers will be blinded to the results of the first reading session.

#### Features

The Index Readers will grade the studies independently on the following features:

- For the PA/AP projection they will assign a KL-grade (0-4). They are instructed that their assigned KL-grade should represent Allocation to Clinical Relevance as described in that section
- For the lateral projection they will rate osteophytes (no/yes) on the proximal and distal patella.
- For both images, they can assign the image as of inadequate quality. They are instructed that the heuristic should be that they would send the patient back for a new radiograph if encountered in daily clinical practice.

### Index test: RBknee

RBknee v2.1 will analyse all studies in the Trial Sample. RBknee takes as input DICOM files encapsulating radiographs of knees in binary format. Output is probabilities of various KL, OARSI grades and tibial eminence osteophytes for PA/AP radiographs and probabilities for presence/absence of proximal and distal osteophytes for lateral radiographs.

### Reader election

One Reference Expert and four Index Readers of different levels of experience and specialty **from each site** will participate in the trial;

- one knee OA expert (Reference Expert, defined above)
- one radiologist in training (Index Reader)
- one radiologist regularly reporting on KOA (Index Reader)
- one knee-specialised orthopaedic surgeon (Index Reader)
- one orthopaedic surgeon in training (Index Reader)

The following information will be collected from both the Reference Experts and the Index Readers:

| Reader information |  |  |
| --- | --- | --- |
| Name | Type | Description |
| ReaderID | Nominal | Unique reader identifier |
| SiteID | Nominal | Clinical site identifier ["BFH","CUB","EMC"] |
| Reader experience | Ordinal | Categorical level of reader experience ["in-training", "specialised", "expert"] |
| Reader specialty | Nominal | Medical specialty of the reader ["radiology", "orthopedic surgery"] |
| Time of evaluation | Continuous | Timestamp of when a study was read by the reader |

## Methods: Data Collection and Management

### Data Collection

Imaging data will be extracted in the DICOM-format from the hospital’s PACS and anonymised using local procedures. Clinical data (age, sex, symptomatic side, radiographic projection, and date of study) will be collected before anonymisation.

The anonymised imaging studies will be uploaded to the online imaging trial platform, ANOVI, to be used for data collection throughout the study. The platform works in the cloud and is compliant to handle pseudonymised (or fully anonymised) imaging data.

### Data management

Each patient will be assigned a unique study ID, e.g., BFH001, EMC001, CUH001, BFH002, EMC002, CUH002 etc. Measurements will be reported in a designated **database**. The clinical data will be manually entered into the database. The feature gradings from ANOVI will automatically be stored and later merged in the **database**.

The measurements that will be collected are:

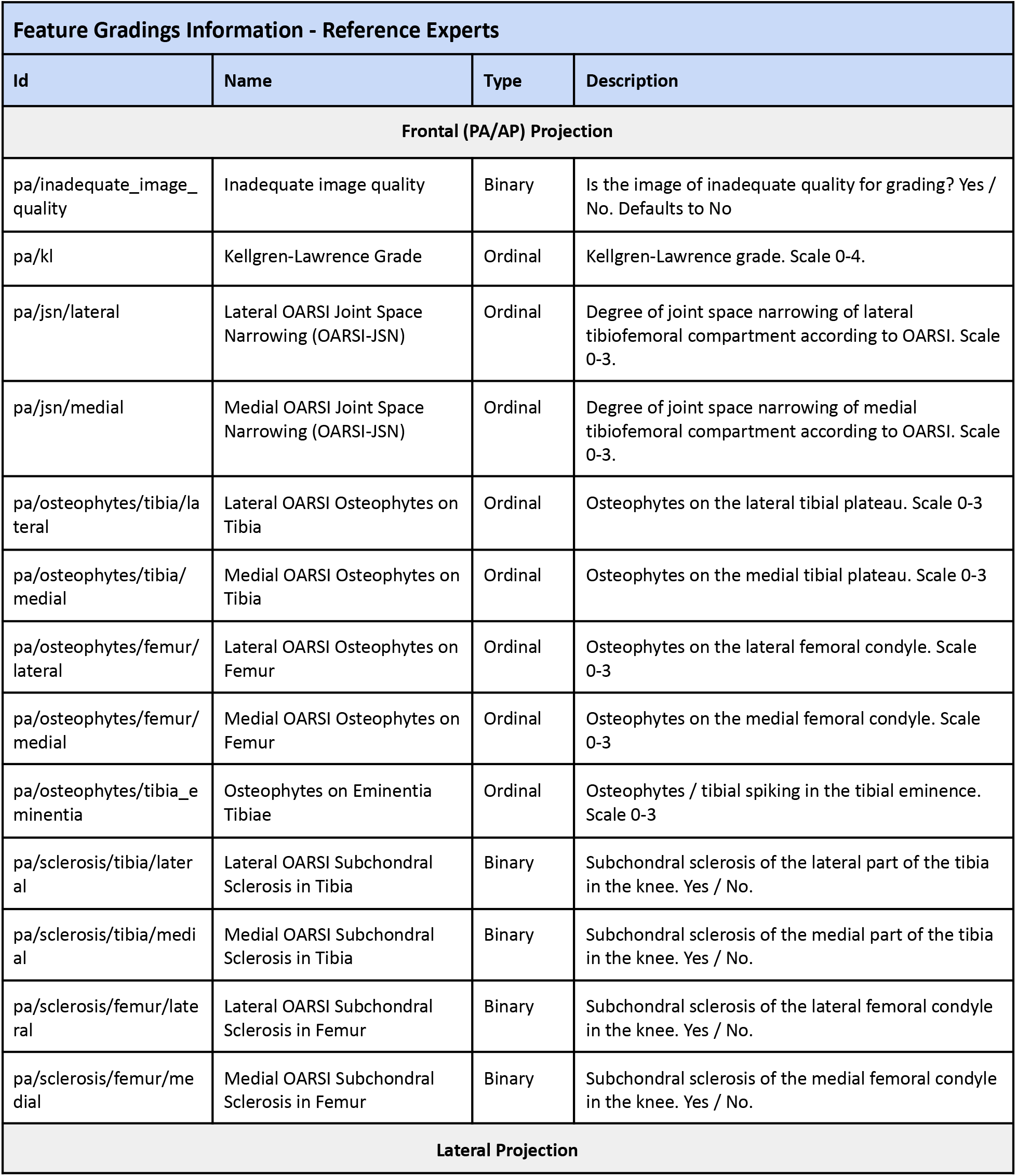
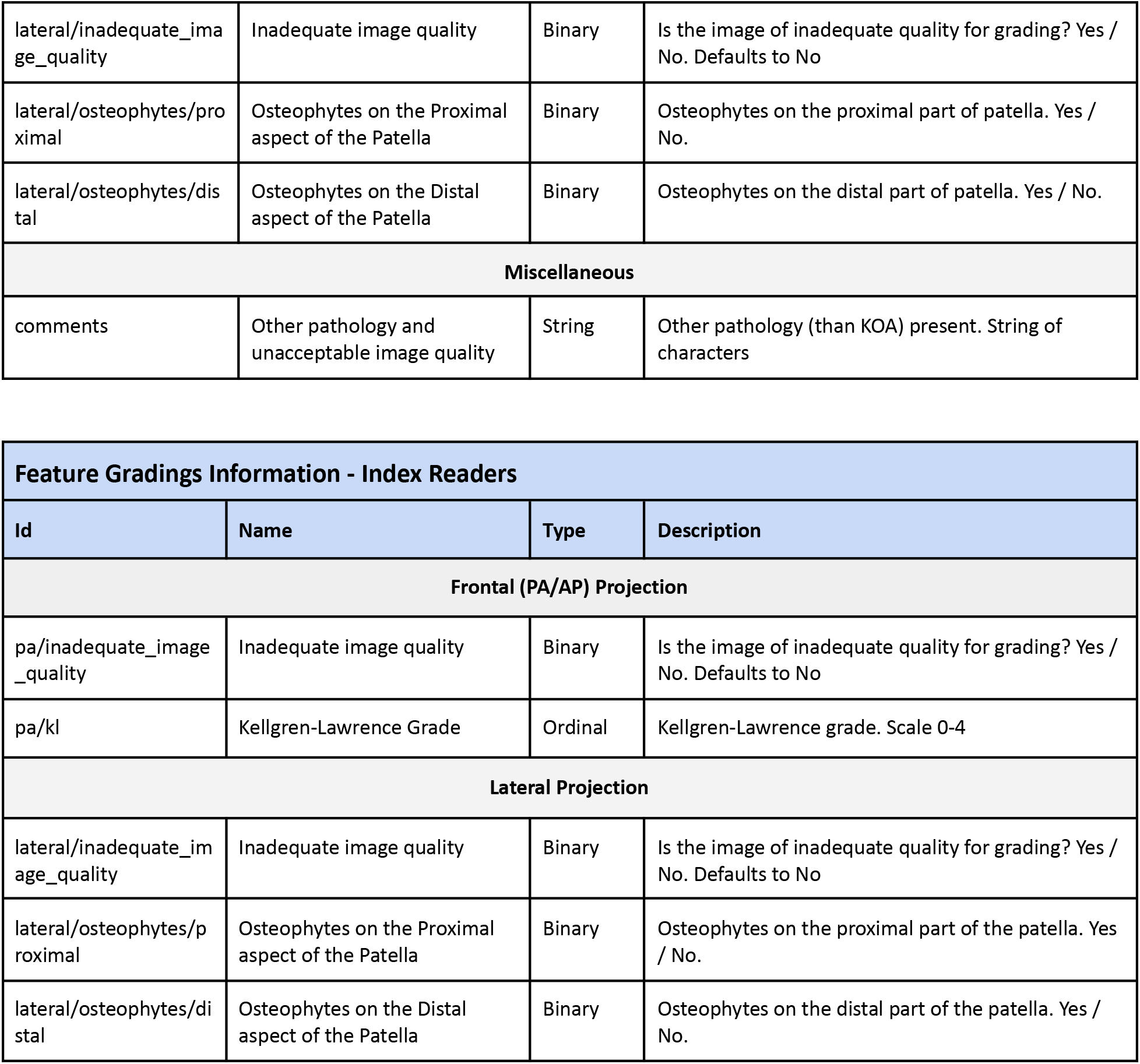

### Statistical methods

#### Index test: Evaluation of Index Readers

Ordinal weighted accuracy for KL-grading will be calculated for all Index Readers without and with decision support from RBknee ^18^.

Balanced accuracy for grading presence/absence of patellar osteophytes on lateral projection images will be calculated for all Index Readers without and with decision support from RBknee.

Quadratic weighted Light’s kappa will be calculated among all Index Readers for KL-grading for the without and the with decision support readings.

For the metrics above, hypothesis testing will be done using permutation tests. In addition, per specialty and per experience level subgroup analyses will also be performed.

#### Index test: Stand alone evaluation of RBknee

Ordinal weighted accuracy will be calculated for RBknee for KL-grading, OARSI-JSN grading, and OARSI-osteophyte grading. Balanced accuracy will be calculated for RBknee for OARSI subchondral sclerosis grading. Hypothesis testing will be done using permutation tests.

## Supporting information

spirit-ai-checklist

## Data Availability

All data produced in the present study are available upon reasonable request to the authors

## Abbreviations

AP: Anterior-posterior
BFH: Bispebjerg and Frederiksberg Hospital
CUB: Charité Universitätsmedizin Berlin
EMC: Erasmus Medical Center
EULAR: European League Against Rheumatism
JSN: Joint-Space Narrowing
KOA: Knee Osteoarthritis
MRMC: Multi-reader, multi-case
OA: Osteoarthritis
OARSI: Osteoarthritis Research Society International
PA: Posterior-anterior

## Ethics and Dissemination

### Regulatory and ethical considerations

The trial is conducted in accordance with the Declaration of Helsinki ^19^ and the General Data Protection Regulation (GDPR) ^20^. Prior to the start of any study activities at each of the sites, local ethics committee opinion and approval from national competent authorities will be collected and, unless the requirement is waived, informed consent will be collected prior to inclusion of patients.

### Protocol amendments

Any significant protocol modifications (e.g. changes in eligibility criteria, outcomes, analysis) must be approved by all investigators and, if necessary, communicated to the respective national ethical authorities.

### Confidentiality

Anonymized data will be entered into the database which allows sharing among the investigators.

### Access to data

All investigators and the sponsor will have access to the final, anonymised trial dataset as recorded in the database.

### Dissemination policy

Findings from this trial are intended for publication in scientific peer-reviewed journals. Furthermore, results will be presented at national and international conferences. All forthcoming studies based on collected data are agreed upon by the trial group. The first author assumes responsibility for all practical issues and the first drafts of all articles. Articles are written and authorship decided according to guidelines by the International Committee of Medical Journal Editors.

